# Reproducibility of the radiosensitivity index and failure of CT radiomics as its surrogate: a public-data study in non-small cell lung cancer

**DOI:** 10.64898/2026.07.20.26358526

**Authors:** Zhe Chen, Rei Gou

## Abstract

**Purpose:** The radiosensitivity index (RSI) and genomic-adjusted radiation dose (GARD) are increasingly treated as quantitative inputs to radiotherapy dose calculation. Two reproducibility issues bear on this use: whether CT radiomics can non-invasively recover RSI, and whether one coefficient of the equation is uniquely specified (printed CDK1 but implemented as PAK2). We examine both on public data.

**Methods:** In GEO GSE103584 RNA-seq (n = 130 non-small cell lung cancer [NSCLC]), we recomputed RSI with the Eschrich 2009 coefficients using CDK1 or PAK2 in the disputed slot and derived GARD under four fixed dose/fractionation schemas. In the paired TCIA NSCLC-Radiogenomics cohort (n = 117), we trained cross-validated Elastic Net and Random Forest models to predict continuous RSI and a median-split RSI label from IBSI-conformant, scanner-corrected CT radiomic features, under a pre-set viability rule.

**Results:** CT radiomics did not recover RSI (Spearman ρ = 0.05 and 0.03; binary AUC = 0.43), below the pre-set viability threshold. Separately, the two probesets listed for the disputed coefficient in the founding paper’s Table 3 both map to PAK2; using the printed CDK1 left rank correlation high (ρ = 0.980) but reclassified 6.2% and 9.2% of patients (median and tertile) and shifted GARD by 4.1–6.3 Gy.

**Conclusions:** CT radiomics is not a viable RSI surrogate in this public cohort, so imaging-GARD should not assume radiomic recovery of RSI. The disputed coefficient resolves to PAK2; implementing the printed CDK1 shifts GARD and reclassifies patients despite high rank correlation. Outcome-directed, dose-adjusted imaging is the more defensible next step.

**Highlights:**

- In 117 paired NSCLC cases, CT radiomics did not recover RSI (ρ≤0.05; AUC 0.43).
- Imaging-GARD assuming a radiomic RSI surrogate is unsupported on public data.
- RSI/GARD enter dose decisions, so each model term must be uniquely specified.
- One RSI coefficient is printed inconsistently (CDK1 vs PAK2); probes support PAK2.
- Using CDK1 not PAK2 shifts GARD 4–6 Gy and reclassifies 6–9% of patients.

**Note added (2026):** After this preprint was first posted, we became aware that the probe-to-gene discrepancy at the −0.0092431 RSI coefficient (printed CDK1, probe-mapped PAK2) had been independently reported by Li M (Int J Radiat Oncol Biol Phys 2025;122:510-513), together with a confirming reply from the original authors (Eschrich and Torres-Roca, ibid. 513-514), who state that their analyses have used PAK2. We defer priority for the gene-identity point to that correspondence. The present work reproduces the finding in NSCLC and additionally quantifies its GARD dose-level consequences and the failure of CT radiomics as an RSI surrogate.

## 1. Introduction

Personalizing radiotherapy dose using tumor biology remains a central aim of medical physics and radiation oncology. The radiosensitivity index (RSI), a 10-gene, rank-based linear model of cellular radiosensitivity, and the genomic-adjusted radiation dose (GARD), which embeds RSI in a linear-quadratic framework, have been proposed as tools to reconcile physical dose with biological effect [1-3]. Downstream work has used RSI/GARD to interpret trial results, propose dose personalization, and motivate imaging surrogates for tissue-free deployment [4-6].

RSI and GARD have moved rapidly from discovery toward implementation: GARD has been associated with radiotherapy benefit across multiple disease sites in a pooled pan-cancer analysis, and RSI-guided prescription dose (RxRSI) has been proposed and entered prospective testing [3,13]. As these constructs approach the clinic, the medical-physics community increasingly treats RSI and GARD as quantitative inputs to dose decisions (on the same footing as dose-volume metrics or the α/β ratio), which raises the bar for their reproducibility and for transparent specification of every term that enters the calculation. A model term whose identity is ambiguous is, in this framing, no different from an undeclared α/β.

In parallel, quantitative imaging (radiomics) has been proposed as a tissue-free route to the same biology, motivated by the frequent absence of sequenceable tissue, particularly after stereotactic body radiation therapy (SBRT), where many early-stage tumors are treated without biopsy. Radiomics is, however, itself sensitive to acquisition, reconstruction and segmentation, and community efforts such as the Image Biomarker Standardization Initiative (IBSI) exist precisely to constrain feature variability [11,14]. Two questions therefore matter independently: whether radiomic features are reproducible, and (assuming they are) whether they in fact encode a molecular radiosensitivity index. The latter has rarely been tested on public, paired imaging-transcriptomic data with a pre-specified success criterion.

Before any quantity may participate in setting a prescription dose, it must satisfy two conditions: it must be obtainable in the patients for whom it is intended, and it must be unambiguously specified. Neither has been examined quantitatively for RSI and GARD, and we test one of each. First, several groups have proposed CT radiomics as a tissue-free surrogate of radiosensitivity or RSI-like phenotypes [5,6]; whether radiomics can recover the genomic RSI itself, the load-bearing step of any “imaging-GARD” construct, has not been cleanly stress-tested on a public, paired imaging-transcriptomic cohort with a pre-committed performance rule. Second, one term of the RSI equation is specified inconsistently: the −0.0092431 coefficient is printed as CDK1 in the founding paper but implemented as PAK2 in much subsequent GARD work [1,2]. These are different transcripts read by different probesets, so the choice changes the numerical definition of the index; the probesets listed for that coefficient in the founding paper’s Table 3 annotate to PAK2. We quantify how often implementing the printed symbol rather than the probe-supported transcript re-ranks patients and shifts GARD, a specification check that should precede any use of the index in a dose calculation.

Here we report a public-data reproducibility study designed for medical physics readership. We (i) test whether CT radiomics recovers genomic RSI in The Cancer Imaging Archive (TCIA) non-small cell lung cancer (NSCLC)-Radiogenomics bridge (n = 117) using cross-validation and a pre-specified viability threshold, and (ii) quantify how the CDK1 vs PAK2 coefficient choice changes RSI and GARD in Gene Expression Omnibus (GEO) GSE103584. The goal is not to propose a new biomarker, but to bound what current RSI/GARD pipelines can and cannot assume.

## 2. Materials and methods

### 2.1. Public datasets

#### Transcriptome

GEO GSE103584 provides processed RNA-seq for the NSCLC Radiogenomics study (gene symbols × samples; n = 130 after download of the GEO supplementary matrix) [7,9]. All ten RSI gene symbols, including both CDK1 and PAK2, were present. Undetected values were set to 0 (lowest expression rank within sample), consistent with a conservative missing-data rule used in our pipeline.

#### Imaging-expression bridge

TCIA NSCLC-Radiogenomics collection provides diagnostic CT and tumor segmentations [7,8,10]. Patients with CT, usable segmentation, and matched RNA-seq formed the bridge cohort (n = 117). This is the same public bridge used previously to train imaging surrogates of molecular phenotypes.

No institutional clinical outcomes were required for the present analyses. All data are publicly redistributable under their respective licenses (GEO; TCIA CC BY 3.0 for the radiogenomics collection as documented by TCIA).

### 2.2. RSI and GARD definitions

Within each sample, the ten RSI genes were ranked by expression (1 = lowest … 10 = highest). RSI was computed with the published coefficients [1]:

*RSI = −0.0098009·AR + 0.0128283·JUN + 0.0254552·STAT1 − 0.0017589·PRKCB − 0.0038171·RELA + 0.1070213·ABL1 − 0.0002509·SUMO1 − 0.0092431·[slot] − 0.0204469·HDAC1 − 0.0441683·IRF1*

Two versions were computed by setting [slot] = CDK1, the symbol printed in the founding equation, or [slot] = PAK2, the symbol used in most subsequent implementations. Higher RSI indicates greater radioresistance. We adopt PAK2 as the primary equation because the two probesets listed for this coefficient in the founding paper’s Table 3 (U24153_at and 205962_at) both annotate to PAK2, and the originating group has since specified the same probeset as PAK2 [23]; the specification and its downstream impact are examined in the Discussion. CDK1 is retained as a comparator because much subsequent work implemented the printed symbol.

GARD was obtained by interpreting RSI as survival fraction at 2 Gy, solving the linear-quadratic model for α with fixed β = 0.05 Gy⁻², then GARD = n·d·(α + β·d) [2,3]. Four illustrative schemas were evaluated without patient-specific DVH: conventional 60 Gy in 30 fractions and 74 Gy in 37 fractions (both 2 Gy per fraction), and SBRT 50 Gy in 5 fractions (10 Gy per fraction) and 48 Gy in 4 fractions (12 Gy per fraction).

### 2.3. Version-impact endpoints (pre-specified)

For RSI(CDK1) vs RSI(PAK2): (1) Pearson and Spearman correlation of continuous RSI; (2) discordance of median high/low labels; (3) discordance of tertile labels (L/M/H); (4) mean absolute GARD difference under each schema, and median-split discordance of GARD. Transcript-level Spearman correlation between CDK1 and PAK2 expression was reported as a sanity check.

### 2.4. CT radiomics extraction

Radiomic features were extracted with PyRadiomics under IBSI-oriented parameters [11,12]: 1 mm isotropic resampling, bin width 25, filters = Original + Laplacian-of-Gaussian (σ = 2, 3, 4) + Wavelet, yielding 1,130 features per tumor in 144 segmented cases (0 extraction failures). The primary surrogate models used Original features only (107 features) after dropping zero-variance columns, to reduce filter-driven overfitting in a small paired cohort. Scanner effects were mitigated by per-Manufacturer z-score of each feature (a deliberately conservative batch correction; more elaborate location-scale harmonization such as ComBat [16] was not pursued because the surrogate was null even after simple correction); Manufacturers with fewer than five patients were pooled as “OTHER”. One manufacturer (GE) accounted for 105/117 patients, so the correction is dominated by within-GE standardization.

### 2.5. Radiomic surrogate models for RSI

Target: genomic RSI computed with the PAK2 slot (primary equation); the CDK1-slot target is reported as a prespecified sensitivity analysis (Supplementary Table S1). Predictors: scanner-harmonized Original radiomic features. Models: Elastic Net (ElasticNetCV, L1 ratios {0.1, 0.5, 0.9}); Random Forest regressor (500 trees); Random Forest classifier for median-split high vs low RSI. Performance was estimated by 5-fold cross-validated out-of-fold predictions (shuffled, fixed seed); the Elastic Net penalty was tuned by inner cross-validation within each training fold, whereas the Random Forest used fixed hyperparameters. Metrics: Pearson r, Spearman ρ (continuous); ROC AUC (binary).

Pre-committed viability rule (locked a priori, before inspecting the harmonized full-cohort test): the surrogate was considered viable only if Spearman ρ ≥ 0.30 or binary AUC ≥ 0.68; otherwise imaging-GARD based on a radiomic RSI is rejected for this cohort. Prespecified robustness analyses (Supplementary Table S1): models re-run without scanner batch correction, and with the CDK1-slot RSI as the target.

### 2.6. Software and reproducibility

Analyses used Python 3.9+ (pandas, NumPy, SciPy, scikit-learn, PyRadiomics). RSI/GARD math and the CDK1/PAK2 comparison scripts are available at https://github.com/zchenmd/rsi-gard-reproducibility and archived at Zenodo (all versions): https://doi.org/10.5281/zenodo.21439665. Random seed = 20260717.

## 3. Results

### 3.1. CT radiomics does not recover genomic RSI

In the paired bridge (n = 117), cross-validated performance was near null (Table 1; Figure 1). The median-split AUC of 0.43 lies within sampling variability of chance at this sample size and does not indicate an inverted association. The pre-committed decision rule was therefore not met. CT radiomics did not provide a usable surrogate of genomic RSI in this public NSCLC cohort.

**Figure 1.** CT radiomics does not recover genomic RSI (TCIA-GEO bridge, n = 117). (A) Out-of-fold radiomics predictions versus true RSI (PAK2, primary equation); Spearman ρ = 0.03 (RF) / 0.05 (Elastic Net); viability gate ρ ≥ 0.30. (B) ROC for median-split high versus low RSI; AUC = 0.43; viability gate AUC ≥ 0.68.

**Table 1.** Performance of radiomic RSI surrogates (pre-committed rule: ρ ≥ 0.30 or AUC ≥ 0.68).

| Task | Model | Metric | Value | Viable? |
| --- | --- | --- | --- | --- |
| Continuous RSI | Elastic Net | Spearman $\rho$ | 0.05 | No |
| Continuous RSI | Random Forest | Spearman $\rho$ | 0.03 | No |
| High/low RSI | Random Forest | AUC | 0.43 | No |

Abbreviations: RSI, radiosensitivity index; AUC, area under the receiver-operating-characteristic curve; $\rho$ , Spearman correlation. Metrics are 5-fold cross-validated in the paired bridge cohort ( $n = 117$ ); the prediction target is the PAK2-slot RSI. The pre-committed viability rule required $\rho \geq 0.30$ or $AUC \geq 0.68$ ; no model met it.

### 3.2. The two candidate transcripts are not interchangeable inputs

In GSE103584 (n = 130), CDK1 was undetected in 11 samples and PAK2 in 0. Spearman correlation between CDK1 and PAK2 expression was ρ = 0.36, confirming that the two transcripts do not track as near-substitutes.

### 3.3. RSI versions correlate highly but reclassify a minority of patients

Continuous RSI(CDK1) vs RSI(PAK2): Pearson r = 0.981, Spearman ρ = 0.980 (both p < 10⁻⁹⁰). Absolute RSI differences: mean |Δ| = 0.056 (max 0.187). Median RSI was 0.506 (CDK1) vs 0.425 (PAK2). Despite high rank correlation, median high/low discordance was 6.2% (8/130 patients), and tertile discordance was 9.2% (12/130). Most swaps were adjacent tertiles (L↔M or M↔H); no L↔H swaps were observed (Table 2; Figure 2).

**Figure 2.** Impact of the disputed coefficient (CDK1 vs PAK2) on continuous RSI (GSE103584, n = 130). (A) RSI computed with the −0.0092431 coefficient assigned to PAK2 (x axis, primary equation) versus CDK1 (y axis); dashed line = identity. Spearman ρ = 0.980; median high/low discordance = 6.2%. (B) Distribution of ΔRSI (CDK1 − PAK2); mean |Δ| = 0.056 (max 0.187).

**Table 2.** Tertile crosstab of RSI(CDK1) vs RSI(PAK2) (n = 130).

| CDK1 \ PAK2 | Low | Mid | High |
| --- | --- | --- | --- |
| Low | 40 | 4 | 0 |
| Mid | 4 | 36 | 2 |
| High | 0 | 2 | 42 |

Abbreviations: RSI, radiosensitivity index; CDK1 and PAK2, the two transcripts reported for the $-0.0092431$ coefficient of the RSI equation. Cells are patient counts ( $n = 130$ ); off-diagonal entries are tertile reclassifications between the two RSI versions.

### 3.4. GARD shifts under fixed schemas

Because GARD is a monotone transform of RSI under fixed (n, d), GARD rank correlation remained high (ρ ≈ 0.98) with the same 6.2% median-split discordance. Mean absolute GARD differences were nonetheless non-trivial (Table 3; Figure 3).

**Figure 3.** Stratification and GARD consequences of the coefficient assignment (CDK1 vs PAK2). (A) Tertile crosstab; rows = CDK1 slot, columns = PAK2 slot (primary equation); tertile discordance = 9.2% (off-diagonal). (B) Mean absolute GARD difference under four fixed dose/fractionation schemas (4.1-6.3 Gy).

**Table 3.**
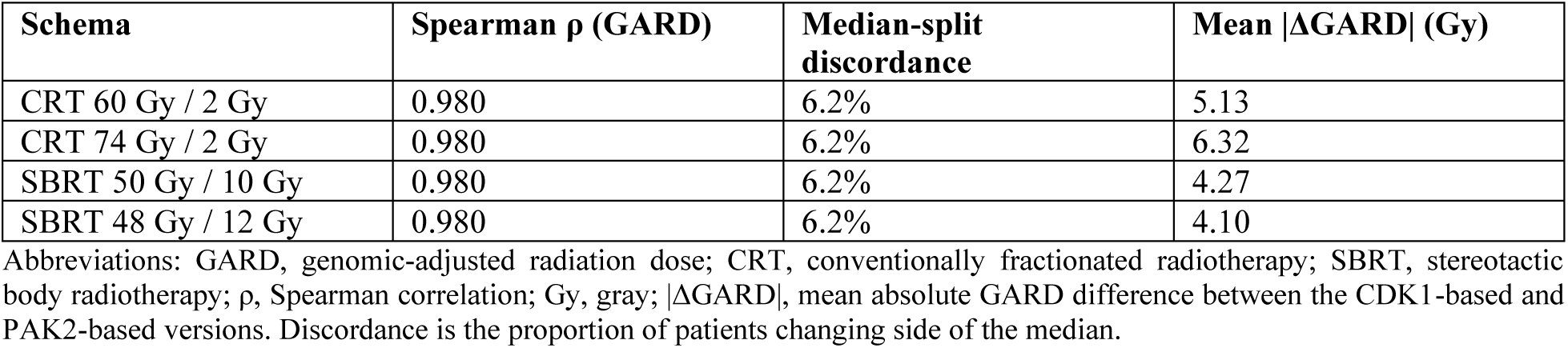
Impact of CDK1 vs PAK2 slot on GARD (n = 130).

## 4. Discussion

### 4.1. Specifying the disputed RSI coefficient in practice

High Spearman correlation between RSI versions can lull analysts into treating CDK1 and PAK2 as “equivalent.” Our results show why that is insufficient for clinical-facing pipelines: a few percent discordance at the median (6.2%) and nearly one in ten patients moving tertile (9.2%), with mean |ΔGARD| of ∼4-6 Gy under standard schemas. For any workflow that thresholds RSI/GARD (e.g., escalate vs de-escalate), the identity of each model term must be declared, frozen, and sensitivity-tested, exactly as dose algorithms declare α/β.

The dissociation between correlation and agreement deserves emphasis because it recurs whenever a candidate index is benchmarked against an established one. A rank correlation of 0.98 describes how faithfully the two versions preserve the ordering of patients; it is insensitive to where patients sit relative to a decision boundary. Clinical use of RSI or GARD, however, proceeds by cutting that ordering at a threshold, so discordance concentrates in the neighbourhood of the cut-point, which is precisely where the residual non-monotonic behaviour is expressed. The structure of the misclassifications supports this interpretation: all tertile changes occurred between adjacent categories and none between the extremes (Table 2), indicating a local perturbation around the boundaries rather than global re-ordering. Reports that a biomarker correlates highly with a reference should therefore be accompanied by the reclassification rate at the thresholds actually used for decisions.

A second observation tempers any reassurance drawn from ρ = 0.98. The two candidate transcripts are not interchangeable at the expression level (ρ = 0.36 between CDK1 and PAK2); the stability of the composite arises because the discordant term contributes one of ten ranks and is diluted by the remaining nine. Apparent robustness of a composite score may therefore reflect averaging rather than unimportance of the substituted component, and cannot be assumed to persist for scores with fewer components or greater weight on the affected term.

The discrepancy can in fact be resolved, and the direction of the resolution determines who is affected. Table 3 of the founding paper labels the hub CDK1 (p34), yet the probe identifiers listed for that row both annotate to PAK2, so any study that selected transcripts by probeset measured PAK2 regardless of the symbol it reported. The mismatch is therefore inert for array-based analyses and becomes consequential only when RSI is reimplemented from gene symbols, which is now the normal case with RNA sequencing. That the two symbols coexisted for more than a decade is unsurprising: both are plausible members of a proliferation-associated signature [21], so neither symbol reads as anomalous on inspection. Plausibility is not numerical equivalence, however, and index-level conclusions have already begun to be drawn from the CDK1 symbol within the RSI panel [22], which the probe evidence does not support. Within the originating group the slot is now specified as PAK2 against the same probeset that the founding paper labelled CDK1 [23], without an accompanying erratum to the symbol (the published correction to the GARD report edits other items but leaves the ten-gene equation unchanged [19]). The reach of the mismatch is illustrated by the most detailed published scrutiny of RSI to date, which enumerates every attainable value of the index and yet lists the slot as CDK1 in its appendix [20]. Papers and software should implement PAK2, state the choice explicitly, and cite the equation source.

### 4.2. Why the radiomics null matters for medical physics

Imaging-GARD constructs require a faithful radiomic surrogate of RSI (or of an equivalent radiosensitivity phenotype). In a fair public test (paired CT and RNA-seq, IBSI-oriented features, scanner batch correction, cross-validated out-of-fold prediction, and a locked viability rule), the surrogate failed (ρ ≤ 0.05; AUC ≈ chance). This does not prove that no imaging feature can ever relate to local control; it does show that assuming radiomics recovers genomic RSI is unsupported. Outcome-directed models that ask whether imaging adds value beyond delivered BED remain a separate, more defensible question and should not be conflated with RSI recovery.

Mechanistically, the null is not surprising. RSI is a within-sample rank combination of ten signalling-hub transcripts; it encodes a specific molecular state that need not manifest as macroscopic CT texture at millimetre resolution. Radiomic features quantify tumour morphology, intensity and spatial heterogeneity, which are shaped by necrosis, atelectasis, vascularity and acquisition, factors only loosely coupled to a ten-gene expression rank. Prior radiogenomic maps have linked imaging phenotypes to broad transcriptional programmes and to prognosis, rather than to any single narrow index [15,17], and previously reported imaging-radiosensitivity associations are typically modest and derived on institutional, non-public bridges [5,6]. Our contribution is to bound this specific claim on public data: even with IBSI-oriented extraction and batch correction, and even reducing to the most reproducible (Original) feature family to limit overfitting, radiomics did not recover genomic RSI. Because one manufacturer dominated the bridge cohort (105/117), the batch correction is conservative by construction; a prespecified re-analysis without harmonization is reported in the Supplementary Material.

Two features of the design warrant explicit statement. First, the viability threshold was fixed before the harmonized full-cohort analysis. Without such a rule, an out-of-fold ρ of 0.15 can be presented as a weak but encouraging association, and the direction of the conclusion becomes a matter of interpretation rather than of data; locking the criterion separates the two. Second, a pre-specified null is not an uninformative result: it bounds what downstream constructs may assume. The proposition falsified here is specifically that CT radiomics recovers the genomic RSI, not the broader proposition that imaging carries no information about radiotherapy outcome. Keeping these distinct matters, because the latter remains open and is the more defensible line of enquiry.

### 4.3. Relation to prior work

Prior imaging-radiosensitivity studies often report modest AUCs for related phenotypes or use institutional, non-public bridges [5,6]. Our contribution is a transparent public benchmark with (i) explicit stress-testing of the coefficient specification and (ii) a negative RSI-surrogate result under pre-committed criteria. Scott et al. and others have used GARD conceptually to interpret dose-escalation trials; those interpretations inherit whatever gene definition and tissue assay were used upstream [3,4]. Reproducibility of the coefficient specification is therefore not pedantic, it is part of the dose-calculation chain. Existing critiques of RSI have addressed its biophysical interpretation, noting that a rank-based linear model can return values outside the interval admissible for a surviving fraction [20]. That line of argument and the present one meet at the same step of the calculation, since GARD derives the radiobiological parameter from RSI read as a surviving fraction, and both bear on whether the index is specified tightly enough to set dose.

### 4.4. Limitations

This study has several limitations. It draws on a single public RNA-seq cohort (GSE103584), so external RNA platforms may shift the absolute RSI scale; GARD was compared under four fixed dose/fractionation schemas rather than patient-specific DVHs; and the radiomic analysis used diagnostic CT segmentations from TCIA rather than planning CT/RTSTRUCT from SBRT. The binary classification used a median split of RSI, and clinically motivated cut-points may differ. As in other cohorts, a minority of RSI values fell outside the interval admissible for a surviving fraction [20]; this affects both RSI versions equally and does not bear on the comparison reported here. Finally, we did not re-analyze published GARD clinical associations under both coefficient versions, which was out of scope and is left as follow-up.

### 4.5. Recommendations for RSI/GARD users

Implement the −0.0092431 slot as PAK2, consistent with the probesets given in the founding paper, and state the choice explicitly with its source. Report version discordance (median and tertile) whenever both versions are computed. Do not treat CT radiomics as a drop-in RSI surrogate without meeting a pre-registered recovery criterion. Finally, when tissue is absent, prefer outcome-directed, dose-adjusted imaging studies (e.g., asking whether imaging adds value beyond the established BED thresholds for SBRT local control [18]) over imaging-to-RSI-to-GARD cascades.

## 5. Conclusions

In public NSCLC data, implementing the printed symbol CDK1 instead of probe-supported PAK2 yields highly correlated RSI scores but reclassifies 6-9% of patients and shifts GARD by several gray on average. Independently, CT radiomics fails to recover genomic RSI in the TCIA-GEO bridge cohort. These results support treating the RSI coefficient specification as a mandatory methods item and discourage imaging-GARD designs that presuppose a radiomic RSI surrogate.

The prospect of prescribing radiotherapy by tumour biology rather than by convention is worth pursuing, and neither result reported here argues against it. What both results argue against is reaching it by shortcut. A dose-determining index whose specification is unsettled is not yet a dose-determining index, and an imaging surrogate that does not recover the quantity it claims to represent cannot carry a dose calculation. Settling that specification, as the α/β ratio is settled, and testing imaging against outcome rather than against RSI, are the conditions under which the original aim remains reachable.

## Declarations

### CRediT authorship contribution statement

Zhe Chen: Conceptualization, Methodology, Software, Formal analysis, Data curation, Visualization, Writing - original draft, Writing - review & editing. Rei Gou: Investigation, Writing - review & editing.

### Declaration of generative AI and AI-assisted technologies in the writing process

During the preparation of this work, the authors used Anthropic Claude (a large language model-based AI assistant) to help write and debug the analysis code and to draft and language-edit the manuscript. The authors reviewed and reproduced all analyses, verified and edited all text, and take full responsibility for the content, results, and conclusions of this article. No AI system is listed as an author.

### Ethics

Public de-identified data only; institutional IRB not required for this analysis.

### Data availability

GSE103584 (GEO); TCIA NSCLC-Radiogenomics. Per-sample RSI(CDK1)/RSI(PAK2) and summary outputs are provided in the code repository.

### Code availability

https://github.com/zchenmd/rsi-gard-reproducibility; Zenodo (all versions): https://doi.org/10.5281/zenodo.21439665

