## Supplementary Table 1 for "Reproducibility of the radiosensitivity index and failure of CT radiomics as its surrogate: a public-data study in non-small cell lung cancer" (from another version of this preprint: version 1, 2026-07-22)

### Supplementary Table S1.

Robustness of the radiomic RSI surrogate to the RSI coefficient choice (CDK1 vs PAK2) and to scanner batch correction (TCIA-GEO bridge, $n$ = 117; Original CT radiomic features). Values are 5-fold cross-validated Spearman ρ (Elastic Net, Random Forest) and median-split ROC AUC (Random Forest). The pre-committed viability rule (ρ ≥ 0.30 or AUC ≥ 0.68) was met in no condition. The PAK2 / with-batch-correction row reproduces the main-text result.

| RSI slot | Scanner batch correction | Elastic Net ρ | Random Forest ρ | Median-split AUC |
|---|---|---|---|---|
| CDK1 | Yes | 0.079 | 0.015 | 0.527 |
| CDK1 | No | 0.116 | 0.018 | 0.499 |
| PAK2 | Yes | 0.050 | 0.032 | 0.434 |
| PAK2 | No | 0.123 | 0.038 | 0.399 |

**Supplementary note.** The radiomic RSI surrogate remained null across all four combinations of RSI coefficient (CDK1 vs PAK2) and scanner batch correction (Table S1). Removing batch correction did not recover signal, Elastic Net ρ rose only marginally (to 0.12) while Random Forest ρ and AUC did not improve (AUC ≤ 0.50), indicating that the correction did not mask a real association. Substituting CDK1 for PAK2 as the prediction target yielded equally null performance. The failure of CT radiomics to recover genomic RSI is therefore not an artefact of the coefficient choice or of scanner harmonization.

### Supplementary data and code

Per-sample RSI(CDK1) and RSI(PAK2), GARD values under each dose/fractionation schema, the tertile crosstab, and the sensitivity results in Table S1 are provided as machine-readable files (rsi_cdk1_vs_pak2_per_sample.csv; gard_cdk1_vs_pak2_schemas.csv; rsi_tertile_crosstab.csv; sensitivity_surrogate.csv).

All analysis code is available at https://github.com/zchenmd/rsi-gard-reproducibility and archived at Zenodo (all versions): https://doi.org/10.5281/zenodo.21439665.
